# Seroprevalence of anti-SARS-CoV-2 IgG antibodies in Kenyan blood donors

**DOI:** 10.1101/2020.07.27.20162693

**Authors:** Sophie Uyoga, Ifedayo M.O. Adetifa, Henry K. Karanja, James Nyagwange, James Tuju, Perpetual Wanjiku, Rashid Aman, Mercy Mwangangi, Patrick Amoth, Kadondi Kasera, Wangari Ng’ang’a, Charles Rombo, Christine Yegon, Khamisi Kithi, Elizabeth Odhiambo, Thomas Rotich, Irene Orgut, Sammy Kihara, Mark Otiende, Christian Bottomley, Zonia N. Mupe, Eunice W. Kagucia, Katherine E Gallagher, Anthony Etyang, Shirine Voller, John N. Gitonga, Daisy Mugo, Charles N. Agoti, Edward Otieno, Leonard Ndwiga, Teresa Lambe, Daniel Wright, Edwine Barasa, Benjamin Tsofa, Philip Bejon, Lynette I. Ochola-Oyier, Ambrose Agweyu, J. Anthony G. Scott, George M. Warimwe

## Abstract

**Background:** There are no data on SARS-CoV-2 seroprevalence in Africa though the COVID-19 epidemic curve and reported mortality differ from patterns seen elsewhere. We estimated the anti-SARS-CoV-2 antibody prevalence among blood donors in Kenya.

**Methods:** We measured anti-SARS-CoV-2 spike IgG prevalence by ELISA on residual blood donor samples obtained between April 30 and June 16, 2020. Assay sensitivity and specificity were 83% (95% CI 59-96%) and 99.0% (95% CI 98.1-99.5%), respectively. National seroprevalence was estimated using Bayesian multilevel regression and post-stratification to account for non-random sampling with respect to age, sex and region, adjusted for assay performance.

**Results:** Complete data were available for 3098 of 3174 donors, aged 15-64 years. By comparison with the Kenyan population, the sample over- represented males (82% versus 49%), adults aged 25-34 years (40% versus 27%) and residents of coastal Counties (49% versus 9%). Crude overall seroprevalence was 5.6% (174/3098). Population-weighted, test- adjusted national seroprevalence was 5.2% (95% CI 3.7– 7.1%). Seroprevalence was highest in the 3 largest urban Counties - Mombasa (9.3% [95% CI 6.4-13.2%)], Nairobi (8.5% [95% CI 4.9-13.5%]) and Kisumu (6.5% [95% CI 3.3-11.2%]).

**Conclusions:** We estimate that 1 in 20 adults in Kenya had SARS-CoV-2 antibodies during the study period. By the median date of our survey, only 2093 COVID-19 cases and 71 deaths had been reported through the national screening system. This contrasts, by several orders of magnitude, with the numbers of cases and deaths reported in parts of Europe and America when seroprevalence was similar.

## Background

Africa accounts for 17% of the global population,^1^ but has reported about 5% of global COVID-19 cases and 2.5% of global COVID-19 deaths to date.^2^ This disparity has been variously attributed to limited capacity for diagnosis, timely implementation of stringent containment measures, a younger population structure^3^ and a predominance of asymptomatic and mild infections.^4^ The first case of COVID-19 in Kenya was detected on March 12, 2020. Within one week the government instituted several containment measures to limit the spread of the virus and the number of fatalities.^5^ While the number of new cases continues to rise (14,168 cases and 250 deaths as at 21^st^ July 2020)^6^, the rate is notably slower than in Wuhan, China or Europe.

The proportion of COVID-19 cases that are asymptomatic varies by setting.^7^ However, in countries like Kenya, with a young population, it is likely to be high.^3^ In addition, testing algorithms have ascertained only a fraction of the symptomatic cases in most countries.^8^ Good public health planning requires information on the potential burden of the pandemic on the healthcare system and the collateral effects of any mitigation measures. In countries affected early, serological surveillance was used to define cumulative incidence. For example, at the release of lockdown in Wuhan, 9.6% of staff resuming work were found to have anti-SARS-CoV-2 antibodies.^9^ At the end of the epidemic wave in Spain, seropositivity was 5.0% in a random population sample of 60,897.^10^ At the tail of the epidemic curve in Geneva, seroprevalence rose over three weeks from 4.8% to 10.9%.^11^ Currently, we do not have any estimates of SARS-CoV-2 seroprevalence in Africa.

Several studies conducted early in the epidemic curve have found seroprevalence substantially below 5%, which emphasises the need for a highly specific antibody assays.^12^ IgG and IgM antibodies rise almost simultaneously within the first 1-3 weeks after symptom onset and 90% of cases are seropositive by 14 days.^12^ IgG responses are weaker in pauci-symptomatic or asymptomatic cases and may wane rapidly.^13^ A wide range of antibody assays have been developed principally against the spike and nucleocapsid proteins of SARS-CoV-2. To facilitate comparisons across studies, the World Health Organisation (WHO) convened a network to share a panel of test sera and evaluate inter-laboratory variation.^14^

Physical distancing and movement restrictions imposed as part of the COVID-19 response make random population sampling impracticable and therefore several countries have monitored seroprevalence in convenience samples from blood transfusion donors^15-17^ or expectant mothers attending ante-natal clinics.^18^ Here we report the results of a pragmatic national serological survey using residual blood samples from transfusion donors across Kenya.

## Methods

### Setting, Participants

This study was carried out at the KEMRI-Wellcome Trust Research Programme (KWTRP) in Kilifi, Kenya in collaboration with the Kenya National Blood Transfusion Service (KNBTS). The KWTRP is the government designated SARS-CoV-2 testing laboratory for the Coastal Region of Kenya.

Anonymised residual donor serum samples used for screening of transfusion transmissible infections were collected at the KNBTS regional centres in 4 sites (Mombasa, Nairobi, Eldoret and Kisumu). The KNTBS guidelines^19^ define eligible blood donors as individuals aged 16-65 years, weighing ≥50kg, with haemoglobin of 12.5g/dl, a normal blood pressure (systolic 120–129 mmHg and diastolic BP of 80–89 mmHg), a pulse rate of 60-100 beats per minute and without any history of illness in the past 6 months. KNBTS generally relies on voluntary non-remunerated blood donors (VNRD) recruited at public blood drives typically located in high schools, colleges and universities. Since September 2019, because of reduced funding, KNBTS has depended increasingly on family replacement donors (FRD) who provide units of blood in compensation for those received by sick relatives.

### Antibody testing using a SARS-CoV-2 Spike Protein ELISA: Assay validation, selection and defining a threshold for seropositivity

We adapted the Krammer Enzyme linked Immunosorbent assay (ELISA) as follows^20^: Nunc MaxiSorp™ flat-bottom 96-well plates (ThermoFisherScientific) were coated with 2µg/ml of whole trimeric spike protein or spike receptor binding domain (RBD) at 37°C for 1h, washed 3 times in wash buffer (0.1% Tween 20 in 1X phosphate buffered saline) and blocked with Blocker™ Casein (ThermoFisherScientific) for 1h at room temperature. Heat-inactivated serum or plasma samples were diluted 1:800 in Blocker™ Casein, added to the spike or RBD-coated plates and incubated for 2h at room temperature. After a further three washes 100µl horseradish peroxidase (HRP) conjugated goat anti-human IgG antibody (KPL-SeraCare) diluted 1:10,000 in wash buffer was added to the plates, incubated for 1h at room temperature, washed 3 times and o-phenylenediamine dihydrochloride (OPD) substrate (Sigma) added for colour development for 10 min. Plates were read on an Infinite® 200 PRO microplate reader (TECAN) at 492 nm and optical density (OD) values for each sample acquired for analysis. The CR3022 monoclonal antibody (mAb) was used as positive control, while a pool of sera from 50 adults sampled pre-COVID-19 pandemic was used as negative control (see article supplement).

We explored different approaches to define seropositivity using well-characterised populations from before and during the COVID-19 pandemic (Table 1, Table S1, Supplementary text). For both the RBD and spike ELISA specificity was higher when using OD ratios (sample OD/negative control OD) rather than the raw sample OD plus 3 standard deviations. By OD ratio, each ELISA correctly classified 901 of the 910 pre-pandemic samples as seronegative and 15 of the 18 known reverse transcriptase polymerase chain reaction (RT-PCR) positive samples as seropositive (Tables 1, S1). However, the spike ELISA detected more seropositive individuals among those who were investigated for SARS-CoV-2 during the pandemic but were RT-PCR negative (Figure S2, panels A & B). Based on these data, we defined anti-SARS-CoV-2 IgG seropositivity as an OD ratio >2 and selected the spike ELISA for this study; the sensitivity and specificity of this threshold was 83% (95% CI: 59-96%) and 99.0% (95% CI 98.1-99.5%), respectively (Table 1, Figure S3 panels A & B).

**Table 1.**
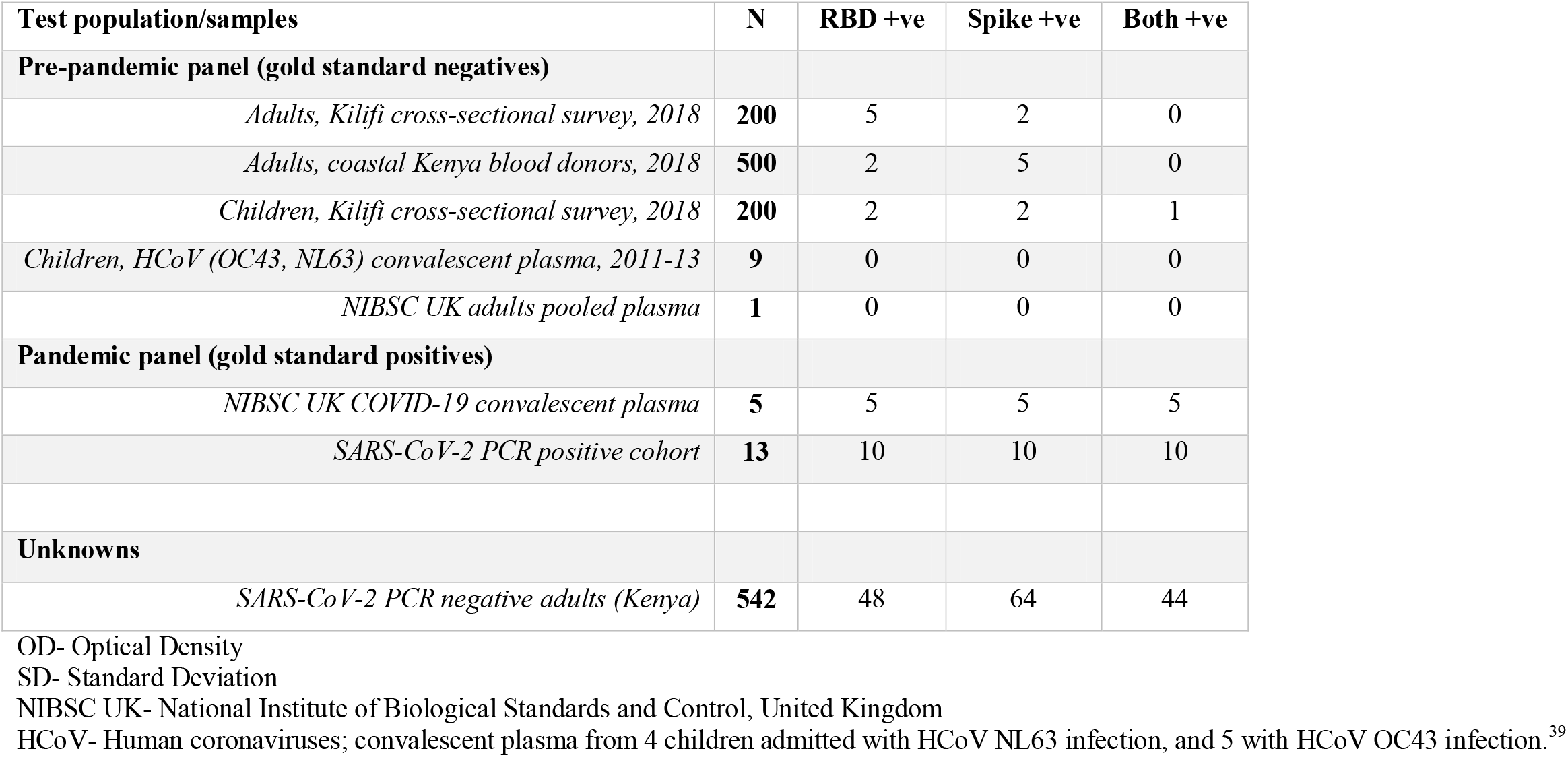
Results for SARS-CoV-2 IgG ELISA validation using sample OD/negative control OD ratio >2 threshold.

### Data Analysis

We tabulated seropositive results by age, sex and residence (county or region). Because the survey was a non-random sample of the Kenyan population, we also calculated standardised prevalence estimates using population data from the Kenya 2019 census. We used 2 methods of standardisation; 1) direct standardisation on the observed prevalence and population weights in 80 region-age-sex strata, and 2) multilevel regression and post-stratification (MLRP) adjusted for the sensitivity and specificity of the assay.^21 22^ To predict stratum prevalence for MLRP we fitted a Bayesian logistic regression model that included sex as a fixed effect and age and region as random effects.^23^ The Bayesian analysis (see Supplement) was done using the rjags package in R version 3.6.1.^24^ We used vague or weakly informative priors for all parameters except the sensitivity (83.0%) and specificity (99.0%) which took bounds from previous studies (62.0%-92.0% and 98.6%-99.8%, respectively).^21^.

We plotted sample distribution and seroprevalence over time to illustrate potential time-trends and tabulated the distribution and seroprevalence across different donor groups (FRDs vs VNRDs) to explore potential biases.

### Ethical Considerations

This study was approved by the Scientific and Ethics Review Unit (SERU) of the Kenya Medical Research Institute (Protocol SSC 3426). Before the blood draw, donors gave individual consent the use of their samples for research.

## Results

### General characteristics of the study population

A total of 3,174 samples were collected between April 30 and June 16, 2020, from individuals aged 15-66 years at four regional centres; approximately half of the samples were drawn in Mombasa, and the remainder were evenly distributed between Nairobi, Kisumu and Eldoret (Figure 1). We excluded 18 duplicate samples, 56 records missing data on age or collection date and 2 records from individuals aged ≥65 years, leaving a total of 3098 (Figure 1).

**Figure 1.**
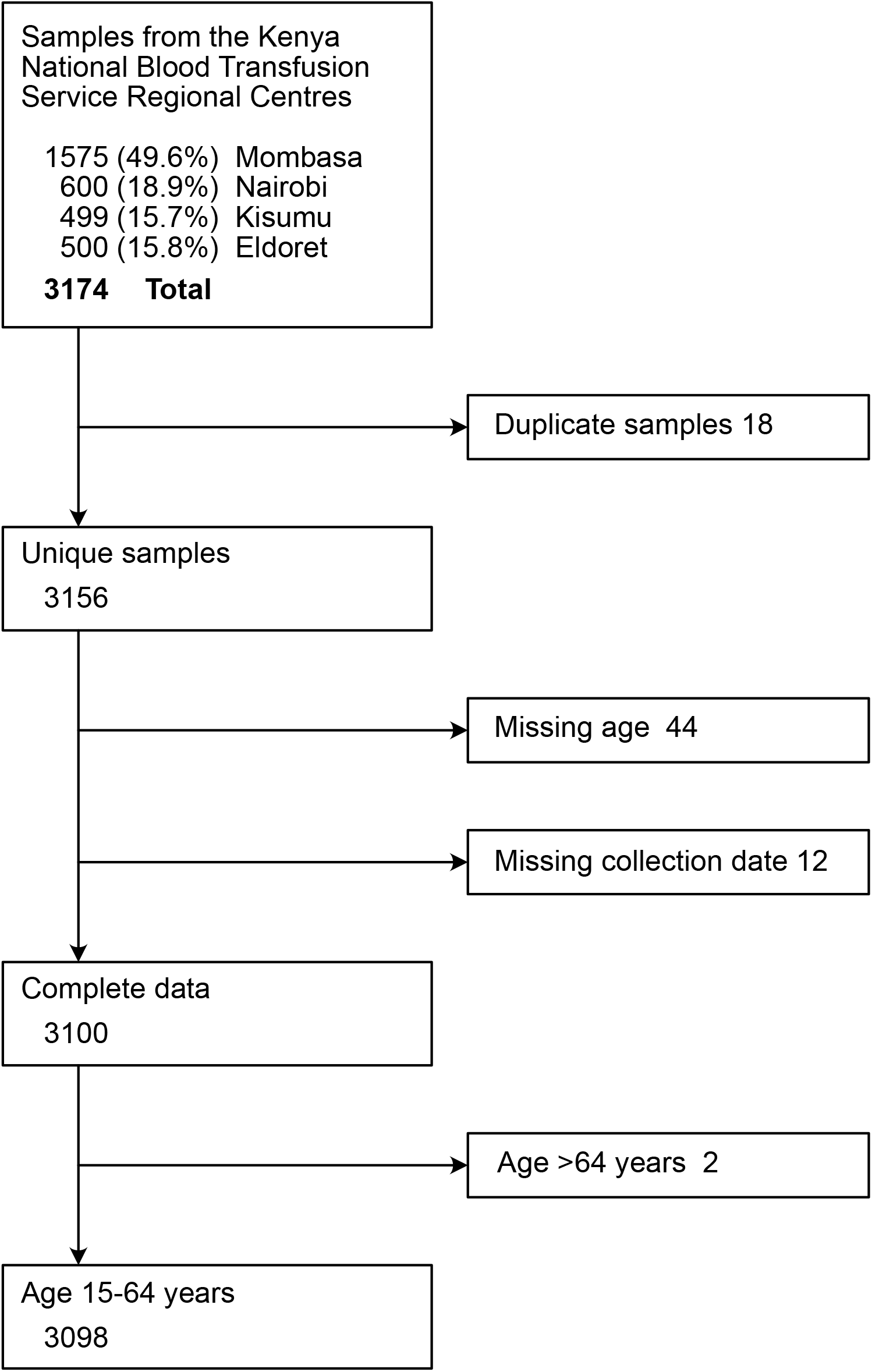
Participant Flow Diagram for SARS-CoV-2 Seroprevalence study of Blood Donors in Kenya.

Compared to the population structure from the 2019 Kenya Population and Housing Census, our participants were predominantly male (82.0% vs 49.3%), had more persons aged 25-34 years (40.1% vs 27.3%) and more residents of coastal Counties (49.2% versus 9.1%, Table 2). In addition, the 15-64-year-old category sampled in our study comprises only 57.1% (27,150,165) of the total population (47,564,296) of Kenya in 2019. Those aged <15 years and ≥65 years make up 39.0% and 3.9% of the total population, respectively.

**Table 2.**
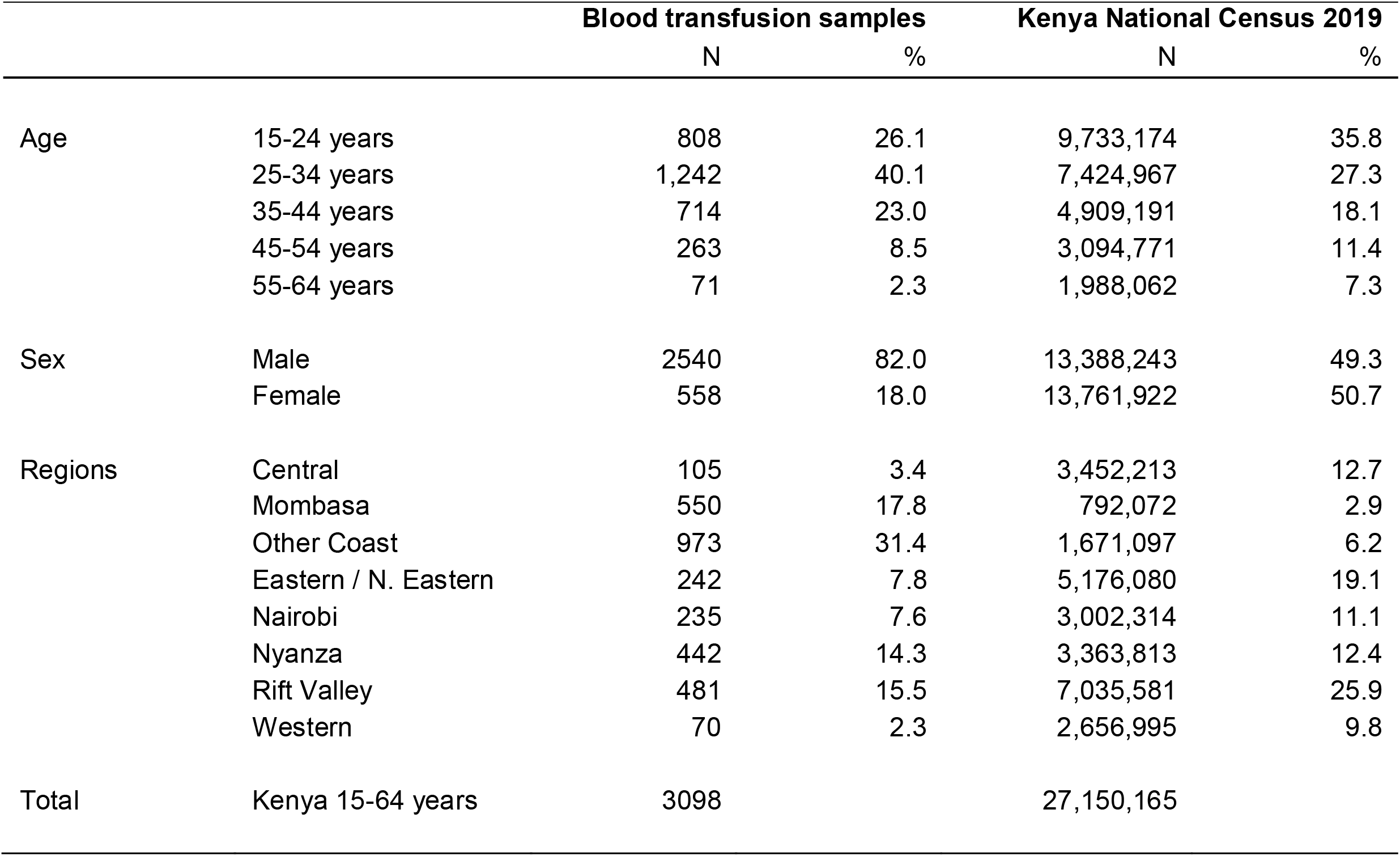
General characteristics of the study population compared to the general population of Kenya.

|  |  | **Blood transfusion samples** |  | **Kenya National Census 2019** |  |
| --- | --- | --- | --- | --- | --- |
|  |  | N | % | N | % |
| Age | 15-24 years | 808 | 26.1 | 9,733,174 | 35.8 |
|  | 25-34 years | 1,242 | 40.1 | 7,424,967 | 27.3 |
|  | 35-44 years | 714 | 23.0 | 4,909,191 | 18.1 |
|  | 45-54 years | 263 | 8.5 | 3,094,771 | 11.4 |
|  | 55-64 years | 71 | 2.3 | 1,988,062 | 7.3 |
| Sex | Male | 2540 | 82.0 | 13,388,243 | 49.3 |
|  | Female | 558 | 18.0 | 13,761,922 | 50.7 |
| Regions | Central | 105 | 3.4 | 3,452,213 | 12.7 |
|  | Mombasa | 550 | 17.8 | 792,072 | 2.9 |
|  | Other Coast | 973 | 31.4 | 1,671,097 | 6.2 |
|  | Eastern / N. Eastern | 242 | 7.8 | 5,176,080 | 19.1 |
|  | Nairobi | 235 | 7.6 | 3,002,314 | 11.1 |
|  | Nyanza | 442 | 14.3 | 3,363,813 | 12.4 |
|  | Rift Valley | 481 | 15.5 | 7,035,581 | 25.9 |
|  | Western | 70 | 2.3 | 2,656,995 | 9.8 |
| Total | Kenya 15-64 years | 3098 |  | 27,150,165 |  |

### Anti-SARS-CoV-2 IgG antibody prevalence in study population

Overall, 174 of 3098 samples were positive for SARS-CoV-2 anti-Spike IgG giving a crude seroprevalence of 5.6% (95% CI 4.8–6.5%). Crude seroprevalence varied by age (p=0.046), ranging between 3.4-7.0% among adults 15-54 years; all 71 donors aged 55-64 years were seronegative (Table 3). Crude seroprevalence did not vary significantly by sex (p=0.50) but did by region, ranging from 1.9% in the Rift Valley region to 10.0% in the Western region (p=0.002, Table 3).

**Table 3.** Crude, population-weighted, and test performance-adjusted SARS-CoV-2 anti-spike protein IgG seroprevalence by participant characteristics and Regions.

| Participant characteristics and Regions |  |  |  |  |  |  |  |  |  |
| --- | --- | --- | --- | --- | --- | --- | --- | --- | --- |
|  | All samples | Seropositive samples | Crude seroprevalence % (95% CI) |  | Kenya population (2019 Census) | Bayesian population-weighted seroprevalence* % (95% CI) |  | Bayesian population-weighted, test-adjusted seroprevalence* % (95% CI) |  |
| Age |  |  |  |  |  |  |  |  |  |
| 15 - 24 years | 808 | 49 | 6.1 | 4.5 - 7.9 | 9,733,174 | 5.1 | 3.7 - 6.9 | 5.3 | 3.5 - 7.8 |
| 25 - 34years | 1242 | 66 | 5.3 | 4.1 - 6.7 | 7,424,967 | 4.9 | 3.6 - 6.4 | 5.1 | 3.5 - 7.3 |
| 35 - 44 years | 714 | 50 | 7.0 | 5.2 - 9.1 | 4,909,191 | 5.9 | 4.3 - 8.1 | 6.2 | 4.1 - 9.3 |
| 45 - 54 years | 263 | 9 | 3.4 | 1.6 - 6.4 | 3,094,771 | 3.8 | 1.9 - 6.0 | 3.9 | 1.5 - 6.7 |
| 55 - 64 years | 71 | 0 | 0 |  | 1,988,062 | 3.4 | 0.7 - 6.2 | 3.7 | 0.7 - 7.0 |
| Sex |  |  |  |  |  |  |  |  |  |
| Male | 2540 | 146 | 5.7 | 4.9 - 6.7 | 13,388,243 | 4.4 | 2.9 - 6.2 | 4.5 | 2.6 - 7.2 |
| Female | 558 | 28 | 5.0 | 3.4 – 7.2 | 13,761,922 | 5.5 | 4.4 - 6.8 | 5.8 | 4.3 - 7.8 |
| Regions |  |  |  |  |  |  |  |  |  |
| Central | 105 | 7 | 6.7 | 2.7 - 13.2 | 3,452,213 | 5.6 | 2.9 - 10.0 | 5.9 | 2.6 - 11.6 |
| Mombasa | 550 | 51 | 9.3 | 7.0 - 12.0 | 792,072 | 8.3 | 6.1 - 10.9 | 9.2 | 6.4 - 12.9 |
| Other Coast | 973 | 39 | 4.0 | 2.9 - 5.4 | 1,671,097 | 3.7 | 2.6 - 5.1 | 3.7 | 2.2 - 5.6 |
| Eastern / N. Eastern | 242 | 11 | 4.5 | 2.3 - 8.0 | 5,176,080 | 4.3 | 2.5 - 7.0 | 4.4 | 2.0 - 7.8 |
| Nairobi | 235 | 21 | 8.9 | 5.6 - 13.3 | 3,002,314 | 7.6 | 4.9 - 11.2 | 8.4 | 4.9 - 13.2 |
| Nyanza | 442 | 30 | 6.8 | 4.6 - 9.5 | 3,363,813 | 6.0 | 4.2 - 8.4 | 6.3 | 3.9 - 9.6 |
| Rift Valley | 481 | 8 | 1.7 | 0.7 - 3.3 | 7,035,581 | 2.1 | 1.1 - 3.6 | 1.9 | 0.7 - 3.8 |
| Western | 70 | 7 | 10.0 | 4.1 - 19.5 | 2,656,995 | 7.0 | 3.5 - 13.1 | 7.7 | 3.3 - 15.5 |
| Total | 3,098 | 174 | 5.6 | 4.8 - 6.5 | 27,150,165 | 4.9 | 3.9 - 6.2 | 5.2 | 3.7 - 7.1 |

\*Re-weighted prevalence estimates based on demographic data from the 2019 Kenya Population and Housing Census.

| Participant Characteristics and Regions |  |  |  |  |  |  |  |  |  |
| --- | --- | --- | --- | --- | --- | --- | --- | --- | --- |
|  | All samples | Seropositive samples | Crude seroprevalence % (95% CI) |  | Kenya population (2019 Census) | Bayesian population-weighted seroprevalence* % (95% CI) |  | Bayesian population-weighted, test-adjusted seroprevalence* % (95% CI) |  |
| Age |  |  |  |  |  |  |  |  |  |
| 15 - 24 years | 808 | 49 | 6.1 | 4.5 - 7.9 | 9,733,174 | 5.1 | 3.7 - 6.9 | 5.3 | 3.5 - 7.8 |
| 25 - 34years | 1242 | 66 | 5.3 | 4.1 - 6.7 | 7,424,967 | 4.9 | 3.6 - 6.4 | 5.1 | 3.5 - 7.3 |
| 35 - 44 years | 714 | 50 | 7.0 | 5.2 - 9.1 | 4,909,191 | 5.9 | 4.3 - 8.1 | 6.2 | 4.1 - 9.3 |
| 45 - 54 years | 263 | 9 | 3.4 | 1.6 - 6.4 | 3,094,771 | 3.8 | 1.9 - 6.0 | 3.9 | 1.5 - 6.7 |
| 55 - 64 years | 71 | 0 | 0 |  | 1,988,062 | 3.4 | 0.7 - 6.2 | 3.7 | 0.7 - 7.0 |
| Sex |  |  |  |  |  |  |  |  |  |
| Male | 2540 | 146 | 5.7 | 4.9 - 6.7 | 13,388,243 | 4.4 | 2.9 - 6.2 | 4.5 | 2.6 - 7.2 |
| Female | 558 | 28 | 5.0 | 3.4 – 7.2 | 13,761,922 | 5.5 | 4.4 - 6.8 | 5.8 | 4.3 - 7.8 |
| Regions |  |  |  |  |  |  |  |  |  |
| Central | 105 | 7 | 6.7 | 2.7 - 13.2 | 3,452,213 | 5.6 | 2.9 - 10.0 | 5.9 | 2.6 - 11.6 |
| Mombasa | 550 | 51 | 9.3 | 7.0 - 12.0 | 792,072 | 8.3 | 6.1 - 10.9 | 9.2 | 6.4 - 12.9 |
| Other Coast | 973 | 39 | 4.0 | 2.9 - 5.4 | 1,671,097 | 3.7 | 2.6 - 5.1 | 3.7 | 2.2 - 5.6 |
| Eastern / N. Eastern | 242 | 11 | 4.5 | 2.3 - 8.0 | 5,176,080 | 4.3 | 2.5 - 7.0 | 4.4 | 2.0 - 7.8 |
| Nairobi | 235 | 21 | 8.9 | 5.6 - 13.3 | 3,002,314 | 7.6 | 4.9 - 11.2 | 8.4 | 4.9 - 13.2 |
| Nyanza | 442 | 30 | 6.8 | 4.6 - 9.5 | 3,363,813 | 6.0 | 4.2 - 8.4 | 6.3 | 3.9 - 9.6 |
| Rift Valley | 481 | 8 | 1.7 | 0.7 - 3.3 | 7,035,581 | 2.1 | 1.1 - 3.6 | 1.9 | 0.7 - 3.8 |
| Western | 70 | 7 | 10.0 | 4.1 - 19.5 | 2,656,995 | 7.0 | 3.5 - 13.1 | 7.7 | 3.3 - 15.5 |
| Total | 3,098 | 174 | 5.6 | 4.8 - 6.5 | 27,150,165 | 4.9 | 3.9 - 6.2 | 5.2 | 3.7 - 7.1 |

\*Re-weighted prevalence estimates based on demographic data from the 2019 Kenya Population and Housing Census.

The Bayesian population-weighted and test-adjusted seroprevalence for Kenya was 5.2% (95% CI 3.7-7.1%, Table 3) and the posterior sensitivity and specificity estimates were 82.5% (95% CI 69.6-91.2%) and 99.2 (95% CI 98.7-99.6%), respectively. Seroprevalence was higher (5.1-6.2%) in the younger age groups (15-44 years) and then declined in the subsequent age groups (45-64 years) but was similar for both sexes. Seroprevalence was highest for Mombasa, Nairobi and the Western region though the number of observations for the Western region was small. The directly-standardised seroprevalence estimates are presented in Table S2 and the directly-standardised national seroprevalence (5.4%) did not differ substantially from the Bayesian population-weighted, test-adjusted estimate. Seroprevalence was also calculated for Counties that had at least 120 donors sampled. The three largest urban Counties of Mombasa, Nairobi, and Kisumu had SARS-CoV-2 seroprevalence of 9.3% (95% CI 6.4-13.2%), 8.5% (95% CI 4.9-13.5%) and 6.5% (95% CI 3.3-11.2%), respectively (Table S3).

The frequency of blood donor sampling and crude seroprevalence estimates increased with time over the 7-week study period (Figure 2). The median sample date was May 30, 2020 while the mid-point of the study was May 24, 2020. We did not adjust for sample date because the period of sampling varied for residents of different counties (Figure 2C) instead we show the variation in crude prevalence over time (Figure 2A).

**Figure 2.**
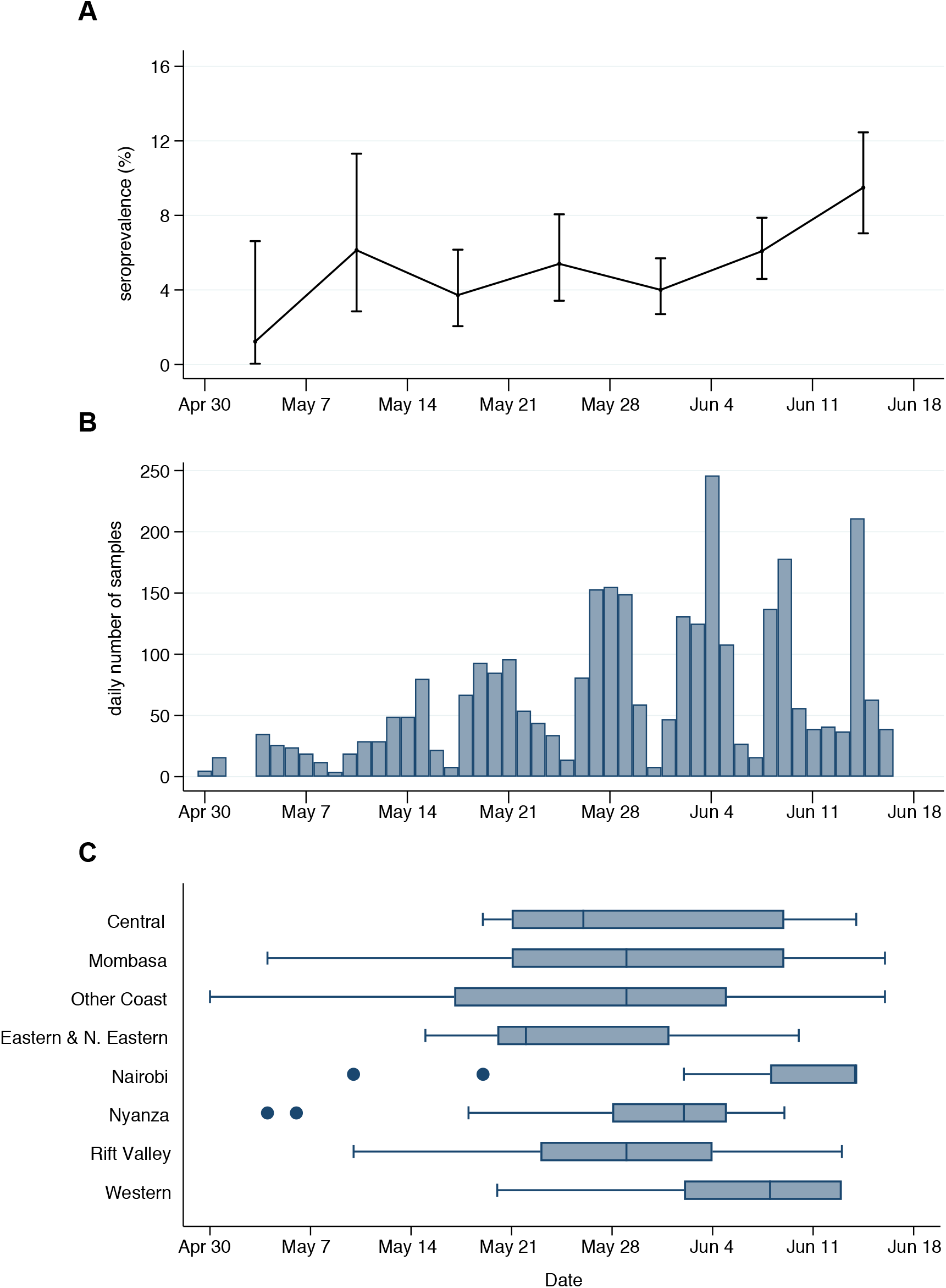
Timeline of sampling for SARS-CoV-2 seroprevalence in blood donors in Kenya. Against the timeline of the sampling period panel A shows the weekly crude seroprevalence, panel B shows the daily frequency of samples collected and panel C shows the temporal distribution of samples by region.

The VNRDs were 7.6% (236/3098) of donors while the remainder were FRDs. The two groups did not differ significantly by age (p=0.15) or sex (p=0.51, Table S4). Crude seroprevalence was 8.5% (20/236) for VNRDs and 5.4% (154/2862) for FRDs. However, the median sample date for VNRDs (June 14, 2020) was two weeks later than the median sample date for FRDs (May 14, 2020).

## Discussion

In this anti-SARS-CoV-2 IgG seroprevalence study of blood donors in Kenya, the crude prevalence was 5.6% and the population-weighted test-adjusted seroprevalence was 5.2%. To the best of our knowledge, this is the first published report of SARS-CoV-2 seroepidemiology in Africa.

Surprisingly, population exposure across the country is considerably higher than previously thought. As expected, seroprevalence was highest in the three urban counties; Mombasa (9.3%), Nairobi (8.5%) and Kisumu (6.5%). Consistent with other studies, seroprevalence did not vary significantly by sex;^25^ it peaked in 35-44-year-olds and was lowest for those ≥45 years, which is also consistent with other studies reporting lower seroprevalence in older adults^10 11^ Antibody responses are related to the severity of disease^25^ and asymptomatic infections, which elicit the least detectable immune responses, are more common in younger populations^4 26^. In older populations, antibody responses may be modified by pre-existing HCoV-elicited immunity^27 28^ and seroprevalence may also be reduced by lower risk of exposure (‘shielding’ at home).

The prevalence of SARS-CoV-2 antibodies in our study is comparable to estimates from large population-based serosurveys in China, Switzerland, Spain and the USA after the initial epidemic peak.^10 11 29 30^ Our results are also comparable to other surveys of blood donors in Brazil^16^, Italy^15^, and many parts of England^17^. Kenya has an estimated population of 53 million in 2020 and 57% of the population is aged 15-64 years. A seroprevalence of 5.2% would therefore suggest approximately 1.6 million infections among 15-64-year olds in Kenya. By the median date of our survey, May 30, 2020, only 2093 cases (of which approximately 90% were asymptomatic) and 71 deaths of all ages had been reported through the national screening system.^31^ The period of sample collection overlapped with the period of government-mandated restrictions; physical distancing measures were first implemented from 20^th^ March 2020.

Anti-SARS-Cov-2 antibody surveys can be used to monitor the spread of the pandemic across the population and to inform mathematical models that predict the course of the epidemic.^32^ Like many other countries, Kenya began easing restriction measures from July 2020 including re-opening of domestic flights which may increase transmission. We intend to monitor SARS-CoV-2 spread by continuous sampling of blood donors throughout the course of the epidemic in Kenya to inform the national response.

Serological surveys can also be used to estimate the state of population immunity. However, the low seroprevalence observed in countries that have experienced a substantial first wave of the epidemic implies that relying on herd immunity is unlikely to terminate the pandemic. At the same time, these observations have provoked a lively debate on alternative mechanisms of immunity to SARS-CoV-2 including cell-mediated immunity^33-35^ and the role of pre-existing HCoV-elicited immunity^27 28^ as well as the function and duration of antibody responses following infection^13 36-38^. Despite our work showing HCoVs circulate in Kenya^39^, we did not identify evidence of cross-reactive antibodies to endemic coronaviruses in the validation of our assay. Other reports suggest that anti-spike antibody responses may be insensitive as a correlate of exposure to the virus or as a correlate of immunity to the diseases.^40^

A key strength of this study is the rigorous validation that included samples from the target population as well as reference plasma from the UK NIBSC as part of a WHO coordinated effort on SARS-CoV-2 seroepidemiology. In addition, we adopted a conservative seropositivity threshold to prioritise specificity over sensitivity. This is relevant in settings like ours, where seroprevalence is under 10% and a similar approach has been implemented by others.^10^

Although blood donors are not representative of the Kenyan population as a whole, we adjusted for bias in the sample structure by standardisation against the age, sex, and regional distribution of the Kenyan population. A substantial proportion (43%) of the population of Kenya are outside the age-range (15-64 years), sampled in this study. The small proportion of older adults in Kenya may be one reason why so few COVID deaths have been observed and the large proportion of children <15 years may explain why so few cases have been observed despite significant evidence of transmission in the population.^3^

Blood donors may also be unrepresentative of the general population because their risk of exposure differs. This bias could work two ways; volunteers are excluded from giving blood if they have been ill during the last two weeks so the sample may underestimate the population SARS-CoV-2 antibody prevalence; on the other hand, people who are shielding at home are unlikely to be captured in our sample leading to an overestimate of seroprevalence. Our exploration of the two distinct populations of blood donors, FRDs and VNRDs, suggests variation in the seroprevalence by donor group but, of note, 92% (2862/3098) of our sample came from the group with lower seroprevalence.

## Conclusions

Our study provides the first national and regional estimates of population exposure to SARS-CoV-2 in an African country. The highest prevalence was seen in the 3 urban counties. The results suggest about 1 in 20 people aged 15-64 years have been exposed to SARS-CoV-2 which is in sharp contrast with the very small numbers of COVID-19 cases and deaths reported during the same period.

## Data Availability

The data and analysis scripts for this manuscript shall be made available at the KWTRP Harvard Dataverse: (https://dataverse.harvard.edu/dataverse/kwtrp). Some of the clinical dataset contains potentially identifying information on participants and is stored under restricted access. Requests for access to the restricted dataset should be made to the Data Governance Committee of the KEMRI-Wellcome Trust Research Programme.

## Acknowledgements

This project was funded by the Wellcome Trust (220991/Z/20/Z; 203077/Z/16/Z). Sophie Uyoga and Charles N. Agoti are funded by DELTAS Africa Initiative [DEL- 15-003], Isabella Ochola-Oyier is funded by a Wellcome Trust Intermediate Fellowship (107568/Z/15/Z), Ambrose Agweyu is funded by a DFID/MRC/NIHR/Wellcome Trust Joint Global Health Trials Award (MR/R006083/1), J. Anthony G Scott is funded by a Wellcome Trust Senior Research Fellowship (214320) and the NIHR Health Protection Research Unit in Immunisation, Ifedayo Adetifa is funded by an MRC/DFID African Research Leader Fellowship (MR/S005293/1) and by the NIHR-MPRU at UCL (grant 2268427 LSHTM). GMW is supported by a fellowship from the Oak Foundation. Charles N. Agoti is funded by the DELTAS Africa Initiative [DEL-15-003], and the Department for International Development and Wellcome (220985/Z/20/Z). We thank Prof Florian Krammer for providing the plasmids used to generate the RBD, spike protein, and CR3022 mAb used in this work. Development of SARS-CoV-2 reagents was partially supported by the NIAID Centres of Excellence for Influenza Research and Surveillance (CEIRS) contract HHSN272201400008C. The COVID-19 convalescent plasma panel (NIBSC 20/118) and research reagent for SARS-CoV-2 Ab (NIBSC 20/130) were obtained from the National Institute for Biological Standards and Control, UK. We thank the blood donors and KNBTS staff who supported this work. We also thank the WHO SOLIDARITY II network for sharing of protocols and for facilitating the development and distribution of control reagents.

This paper has been published with the permission of the Director, Kenya Medical Research Institute.

